# Denial of Access as an Unmeasured Outcome in Nonprescription Statin Self-Selection Studies

**DOI:** 10.64898/2026.09.06.26362346

**Authors:** Kyle P. Molinari

**Affiliations:** Krafted Therapeutics, Inc., Havertown, Pennsylvania, USA

**Keywords:** nonprescription drugs, self-selection studies, statins, sensitivity and specificity, decision curve analysis, additional condition for nonprescription use (ACNU)

## Abstract

**Background:** Five nonprescription statin attempts have failed; a sixth attempt, under the FDA’s 2025-issued Additional Condition for Nonprescription Use (ACNU) pathway, is under way. Self-selection evidence is evaluated by whether ineligible consumers are excluded; how often eligible consumers are denied has never been reported.

**Objective:** To quantify denial of access to eligible consumers across the published self-selection record.

**Methods:** Secondary analysis of the three published United States self-selection studies (SELECT, CREST, TACTiC), the FDA guidance, and four advisory committees’ questions and votes. We recomputed diagnostic accuracy from published cross-tabulations, derived TACTiC’s prespecified unmitigated endpoints, and compared access strategies by decision curve analysis.

**Results:** The guidance’s endpoints contain no measure of denial, and every committee question addressing a selection error concerns admission of the ineligible. CREST’s sensitivity was 59.0% (95% CI 42.1%–74.4%), 29.5 percentage points below its reported positive predictive value; 41% of clinician-eligible participants were not approved. TACTiC’s reference standard reached only application-approved consumers; none of the 10,332 rejected was assessed; sensitivity is inestimable and its prespecified unmitigated endpoints, 84.2% and 80.9% against the published mitigated 90.7% and 98.1%, remain unpublished. Decision curve analysis favors the application over both alternatives across reasonable thresholds at the sample prevalence.

**Conclusions:** For nonprescription statin applications, the self-selection evidence framework counts errors of admission and leaves errors of denial unmeasured; its public health objective, fewer eligible adults untreated, cannot be evaluated on evidence counting only admissions. Four design and reporting changes, none requiring new methodology, would let the next application measure it.

## 1. Introduction

Statin therapy reduces cardiovascular morbidity and mortality across a wide range of baseline risk [1], and eligibility criteria for it are defined by widely disseminated clinical guidelines [2]. Many adults who meet the criteria are nonetheless untreated. In the Provider Assessment of Lipid Management (PALM) registry, which sampled 5,905 statin-eligible primary and secondary prevention patients across 138 of the registry’s 140 United States practices, 74.7% of eligible adults were receiving a statin and 42.4% were receiving guideline-recommended intensity; among primary prevention patients, 63.4% were on a statin [3]. These figures describe patients already established in care, and are therefore a conservative estimate of undertreatment in the broader eligible population [3, 4]. The shortfall extends beyond initiation: among patients prescribed statins in primary care, low adherence and failure to reach guideline lipid-lowering targets are common [5]. Expanded consumer access has been proposed as one response. Yet across the 45 prescription-to-nonprescription switches the US Food and Drug Administration (FDA) approved between 2001 and 2025, spanning 30 active ingredients, no cardiovascular agent appears [6]. The therapeutic class with the largest evidence base in preventive cardiology has never achieved nonprescription status in the United States.

Five attempts at an over-the-counter (OTC) statin by four sponsors have failed, and the reason changed across them [7, 8]. In July 2000 a joint meeting of the Nonprescription Drugs Advisory Committee and the Endocrinologic and Metabolic Drugs Advisory Committee considered Merck’s lovastatin 10 mg and, the next day, Bristol-Myers Squibb’s pravastatin 10 mg. On both days the committee agreed unanimously that a clinical benefit defined as lowering of low-density lipoprotein cholesterol (LDL-C) had been demonstrated, rejected almost as decisively a benefit defined as reduction of cardiovascular events, and voted the application down [9, 10]; the deliberations turned on the existence and definition of clinical benefit, and self-selection appeared only as a sub-consideration within a single question at each meeting [9, 10]. The third attempt, lovastatin 20 mg supported by the Consumer Use Study of Over-the-Counter Lovastatin (CUSTOM), in which 84% of 3,316 consumers evaluating the product in storefront settings made appropriate initial use decisions [11], was heard in January 2005. That committee accepted the drug, its dose, and its safety for nonprescription use, but voted against the proposition that the frequency of appropriate self-selection supported safe and effective nonprescription use, and unanimously that post-initiation self-management raised concerns [12]. The fourth, supported by the Self Evaluation of Lovastatin to Enhance Cholesterol Treatment (SELECT) study of 1,326 consumers [13], was rejected in December 2007 on the same self-selection grounds [14]. The fifth, Pfizer’s actual use trial of atorvastatin 10 mg in a simulated OTC environment, did not meet its objectives for consumers’ compliance with LDL-C testing and action on the result, and was discontinued [7, 15]. The binding constraint moved: first the drug, then the consumer’s initial decision, then the consumer’s behavior over time.

The FDA’s 2012 public hearing on using innovative technologies and other conditions of safe use sought to expand which drug products can be considered nonprescription, and eventually led to a 2018 draft guidance on innovative approaches for nonprescription drug products, the initial development of a category in which a sponsor may impose conditions beyond labeling to support appropriate consumer use [16, 17]. The final rule establishing nonprescription drug products with an additional condition for nonprescription use (ACNU) was issued in December 2024 and, after two delays of its effective date, took effect on May 27, 2025 [18–20]; a Center for Drug

Evaluation and Research procedures manual on prescription-to-nonprescription switches followed in December 2025 [21]. Technology-assisted self-selection, in which a software application rather than a printed label determines eligibility, is the form of ACNU proposed to answer the objection on which the earlier applications failed, and a sixth attempt, AstraZeneca’s rosuvastatin 5 mg OTC program, is built on it [7, 22]. Two studies constitute its evidence base: the Consumer Rosuvastatin Electronic Self-Selection Trial (CREST), which compared 500 participants’ self-assessments against blinded clinician assessment [7], and the Technology-Assisted Cholesterol Trial in Consumers (TACTiC), a six-month self-selection and actual use study of 1,196 participants [22].

This secondary analysis examines how the 2013 FDA guidance on self-selection studies [23] operationalizes correct self-selection, and follows that choice through CREST and TACTiC, asking what the resulting endpoints measure and what they leave unmeasured. Its focus is the outcome the framework does not record: the eligible consumer who is denied access.

## 2. Materials and Methods

This study is a secondary analysis of publicly available published data and regulatory documents. No new data were collected.

### 2.1 Design and data sources

This analysis centers on AstraZeneca’s TACTiC study [22] and the CREST study that preceded it [7]; earlier statin programs enter as comparators in the endpoint analysis and the Discussion. Source material comprised three classes. The first comprised the materials of the two studies mentioned above: the pivotal publications [7, 22], the published supplemental material for TACTiC [22], its redacted clinical study protocol [8] and statistical analysis plan [24] released by AstraZeneca, and the ClinicalTrials.gov results record for TACTiC (NCT04964544; accessed August 22, 2026) [25]. Both the protocol and the statistical analysis plan are also included in the publication’s supplemental appendix [22]. The second comprised material on statin use and on the statin self-selection programs that preceded CREST and TACTiC: the guideline and registry literature on statin eligibility, treatment, and adherence [1–5], the CUSTOM [11] and SELECT [13] publications. The third comprised the regulatory record: the FDA guidance documents on self-selection studies (April 2013) [23] and label comprehension studies (August 2010) [26]; the Federal Register notices establishing the additional conditions for nonprescription use pathway, comprising the 2012 public hearing notice (77 FR 12059) [16], the 2018 draft guidance notice (83 FR 33938) [17], the final rule (89 FR 105288) [18] and its delays of effective date [19, 20]; the manual of policies and procedures governing prescription-to-nonprescription switch applications (CDER MAPP 5200.11) [21]; and the advisory committee record for the four applications heard before a joint meeting of the Nonprescription Drugs Advisory Committee and a therapeutic committee, comprising the agendas, minutes, and questions to the committee for the meetings of July 13 and 14, 2000 (docket 3622) [9, 10], January 13 and 14, 2005 (docket 2005-4086) [12], and December 13, 2007 (docket 2007-4331) [14], together with the December 2007 briefing materials [27] and the minutes and transcript of the Nonprescription Drugs Advisory Committee meeting of September 25, 2006 (docket 2006-4230) [28], cited in the 2013 guidance as the source of the advice it incorporates [23].

### 2.2 Endpoint architecture analysis

Section III.C.1 of the FDA’s 2013 self-selection guidance [23] presents a two-by-two table of possible self-selection outcomes (Figure 1), with cells labeled A through D: selection by a consumer for whom the product is appropriate (A), selection by a consumer for whom it is not (B), non-selection by a consumer for whom it is appropriate (C), and non-selection by a consumer for whom it is not (D). The guidance offers illustrative candidate primary endpoints built from those cells. We translated the table into standard diagnostic accuracy terms, treating the consumer’s self-selection decision as the index test and the clinician’s independent assessment as the reference standard; cell B is then a false positive and cell C a false negative. When applying that translation to the guidance’s illustrative endpoints, it gives each its standard name; the guidance’s A/(A+B), for example, is positive predictive value. Applied to the endpoints reported by the three published consumer self-selection studies of nonprescription statins conducted to date in the United States (SELECT [13], CREST [7], and TACTiC [22]), it identifies which measures were reported and which were not. Applied to the questions FDA posed to advisory committees at the meetings of July 2000, January 2005, and December 2007, it classifies each question by the error it concerns: inappropriate access to the product by a consumer for whom it was not indicated (cell B), or denial of access to a consumer for whom it was indicated (cell C).

**Figure 1.**
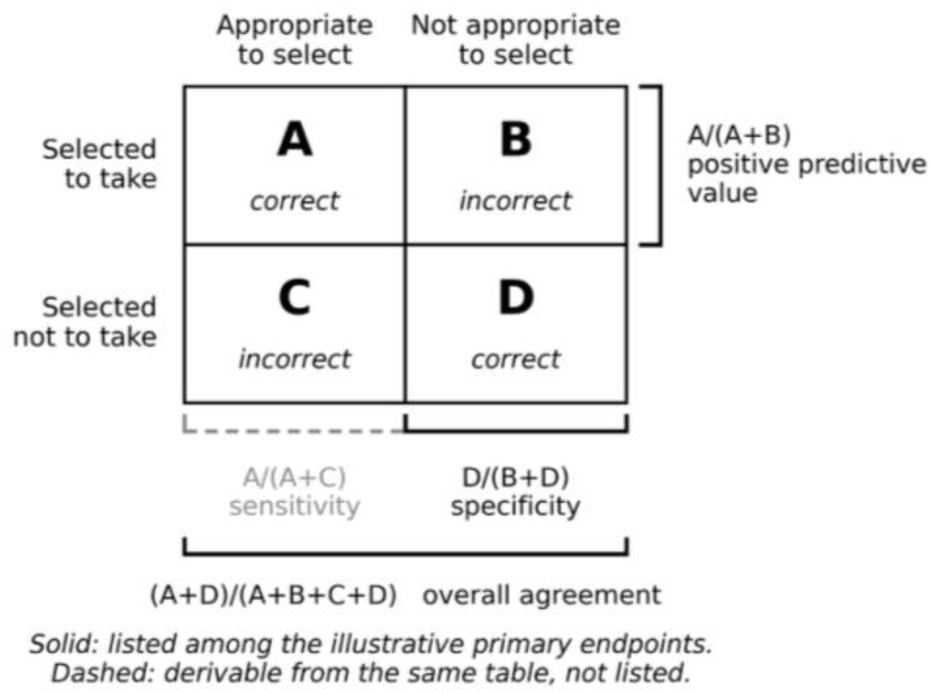
Outcome table presented in Section III.C.1 of the FDA guidance on self-selection studies, with the measures derivable from it Brackets indicate the cells each measure spans. Solid brackets denote measures listed among the guidance’s illustrative primary endpoints; the dashed bracket denotes sensitivity, derivable from the same table but not listed.

### 2.3 Accuracy reanalysis of CREST

In CREST, participants were recruited by digital advertising, traditional media, and community outreach and screened through a study call center; each completed the self-selection assessment unaided on a web-based application at home or elsewhere outside a research facility, and the outcome the application returned was not disclosed to them [7]. Each then attended a scheduled visit at a research site, where staff verified the laboratory and blood pressure values entered, obtaining a fingerstick lipid panel and a blood pressure measurement where verification could not be produced, and forwarded the verified information to a telemedicine clinician who was unaware of the participant’s entries and who completed an independent assessment in the same application; that assessment is the reference standard, and, of 1,063 consumers sent the application link, 500 completed both [7]. CREST preceded TACTiC as its proof of concept; its enrollment period is not reported in the publication [7, 22].

CREST reports, for all 500 participants who completed both assessments, a three-by-three cross-tabulation pairing each participant’s outcome (“OK to use,” “ask a doctor,” or “not right for you”) with the outcome returned by the clinician’s independent assessment [7]. Reducing it to the guidance’s two-by-two form requires a rule for “ask a doctor.” We defined the index test as positive when the participant self-selected “OK to use,” the only outcome granting access, and the reference standard as positive when the clinician’s assessment returned “OK to use,” all other clinician outcomes counting as ineligible; clinician-eligible participants who received “ask a doctor” were counted in the primary analysis as eligible participants not granted access, and excluded in a secondary analysis (Supplementary Methods S1) [7].

Sensitivity, specificity, predictive values, overall agreement, and a null classifier returning “not right for you” to every participant were computed from the pre-mitigation cross-tabulation with two-sided 95% Clopper-Pearson exact intervals [29]; the interval for the difference between positive predictive value and sensitivity used a multinomial bootstrap of the cross-tabulation (200,000 resamples, percentile method) [30]. The single mitigated outcome in CREST affects only negative predictive value, which is reported both ways (Supplementary Methods S1).

### 2.4 Recomputation of the TACTiC co-primary endpoints

TACTiC was conducted in United States markets between July 2021 and March 2023 (NCT04964544) [22, 25], entirely by virtual visit with no in-person contact: consumers who responded to advertising and passed a prescreen were emailed a link and completed the web application unaided, and only those receiving an “OK to use” or an “ask a doctor” outcome were eligible to consent, of whom 1,196 did [22]. At the first virtual visit a Central Medical Operations Group clinician (a registered nurse, nurse practitioner, physician assistant, or physician), blinded to the participant’s application entries, obtained a targeted medical and medication history and completed an independent application assessment using only source-verified laboratory and blood pressure data; the same blinded procedure was repeated at the second virtual visit to determine the final use outcome [8, 22, 24].

TACTiC’s co-primary endpoints are the proportion of participants with a correct initial self-selection outcome and the proportion with a correct final use outcome, each with a prespecified success criterion requiring the lower bound of its 95% confidence interval to exceed 85% [8, 22, 24]. The endpoints count a participant as correct when the participant’s application outcome and the clinician’s independent assessment agree, and also when the two disagree, but a clinical review independent of the assessing clinician determines that the participant’s outcome was nonetheless appropriate; these reclassified cases are termed mitigated outcomes [8, 22, 24]. The first endpoint is assessed in all 1,196 consented participants and the second in the 1,154 who completed the final use assessment or had sufficient information to determine it [22, 24]. Component counts for each endpoint, including the mitigated component, are reported in the publication’s primary outcomes table, and the composition of the mitigated component in its supplemental tables [22].

The unmitigated figures are not reported in the publication, its supplemental material, the protocol, the statistical analysis plan, or the ClinicalTrials.gov results record [8, 22, 24, 25], and were not identified in any other public source; they are identified as derived wherever they appear in this manuscript.

Each unmitigated endpoint is derived by excluding the mitigated participants from the numerator; two-sided 95% Clopper-Pearson exact intervals are calculated by the method prespecified in the statistical analysis plan [24], comparing each lower bound with the prespecified 85% criterion [8, 24], and determining the minimum number of mitigations that would need to be restored for each endpoint to meet it. As a check that this interval method matches the study’s statistical analysis plan, the endpoints were recomputed as reported and reproduced the published intervals exactly (Supplementary Methods S2) [22].

### 2.5 Decision curve analysis

The net benefit of screening by the CREST application was evaluated against the two default comparators of decision curve analysis, which bound the space of access policies: granting every consumer access without screening, and granting none [31, 32]. The analysis is possible only for CREST, because net benefit requires the reference standard on both the consumers the application approves and those it rejects, and TACTiC applied it only to the approved (Section 3.3). Net benefit was calculated as

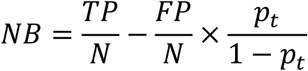

where TP and FP are the clinician-eligible and clinician-ineligible consumers approved by a strategy (23 and 3 for the application, from Table 2 Panel A; 39 and 461 under unscreened access), N the sample size, and p_t_ the threshold probability, the probability of guideline eligibility at which taking the product first becomes worthwhile to the consumer (not the ten-year cardiovascular risk threshold of the treatment guidelines). Curves were computed across all thresholds, and primary interpretation was restricted to a reasonable range of 5% to 20% specified on clinical grounds, following published recommendations [31, 32]; the justification of the range and the reading of a threshold as an exchange rate between the two errors are given in Supplementary Methods S3. The thresholds at which the application ceases to outperform each comparator and report their position relative to that range were identified.

**Table 1.**
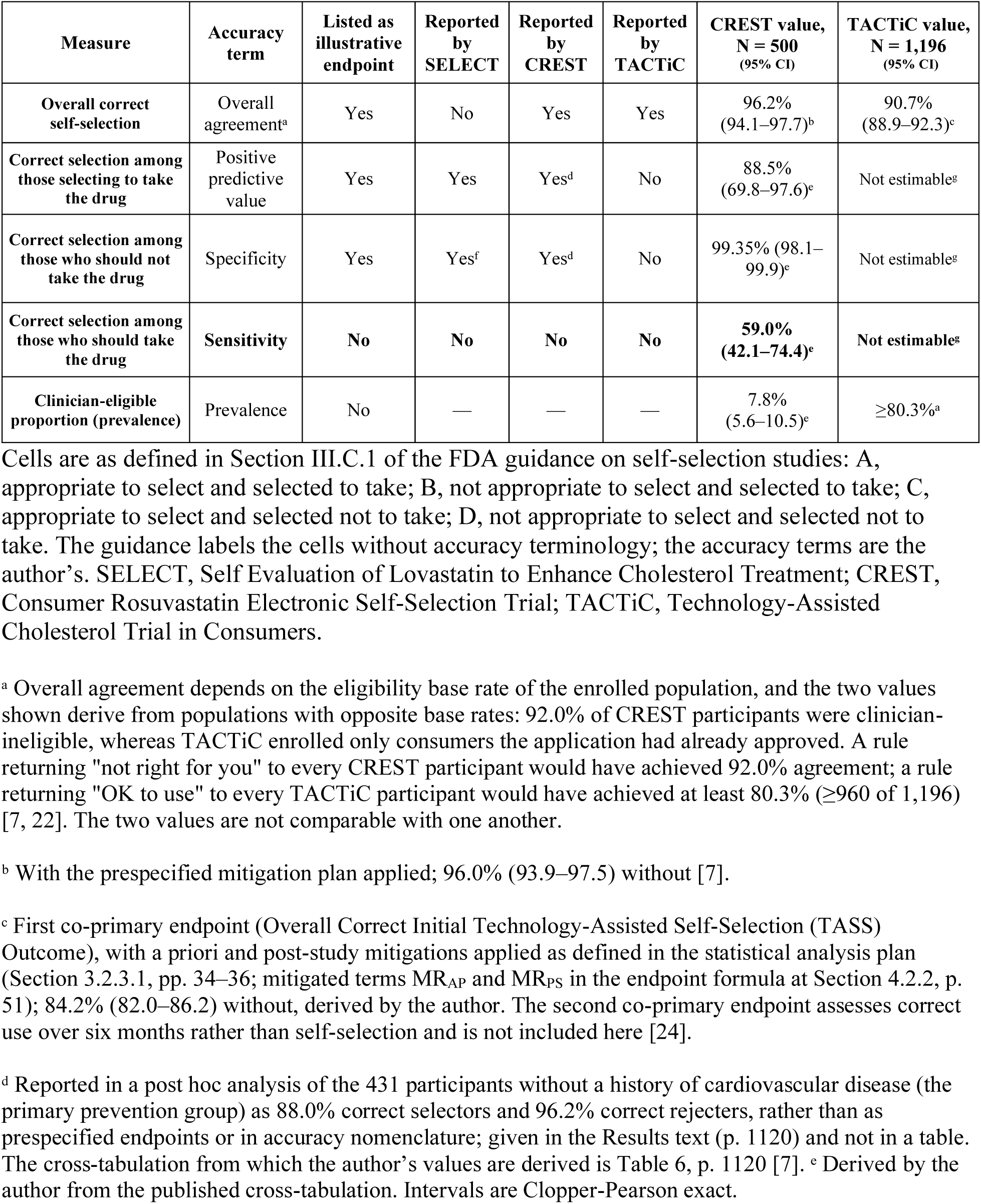
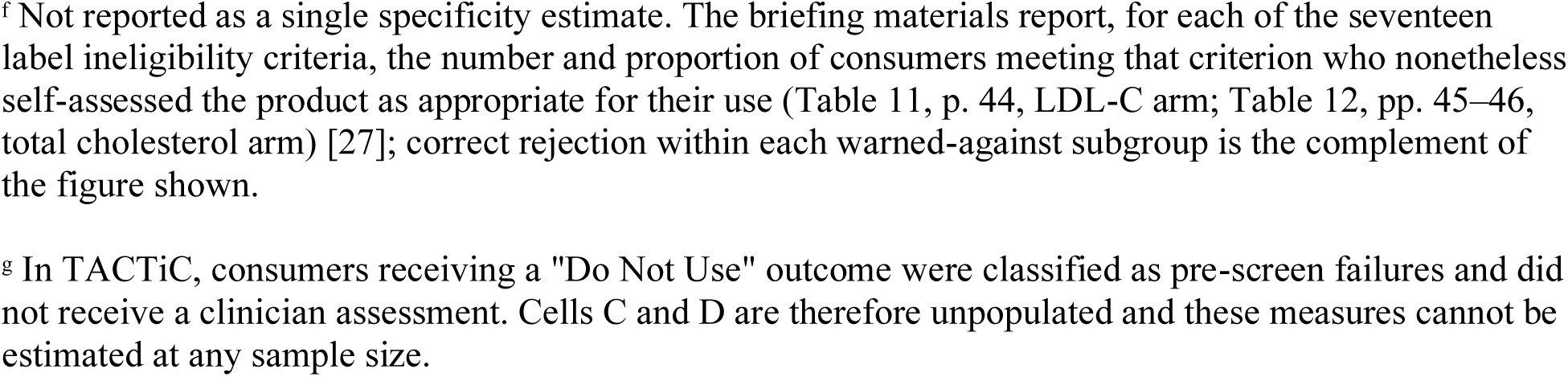
Self-selection accuracy measures derivable from the two-by-two outcome table in FDA guidance: whether each is listed among the guidance’s illustrative primary endpoints, which published nonprescription statin self-selection studies report each, and the values obtained in CREST and TACTiC [7, 13, 22, 27]. Cells are as defined in Section III.C.1 of the FDA guidance on self-selection studies: A, appropriate to select and selected to take; B, not appropriate to select and selected to take; C, appropriate to select and selected not to take; D, not appropriate to select and selected not to take. The guidance labels the cells without accuracy terminology; the accuracy terms are the author’s. SELECT, Self Evaluation of Lovastatin to Enhance Cholesterol Treatment; CREST, Consumer Rosuvastatin Electronic Self-Selection Trial; TACTiC, Technology-Assisted Cholesterol Trial in Consumers. ^a^ Overall agreement depends on the eligibility base rate of the enrolled population, and the two values shown derive from populations with opposite base rates: 92.0% of CREST participants were clinician-ineligible, whereas TACTiC enrolled only consumers the application had already approved. A rule returning “not right for you” to every CREST participant would have achieved 92.0% agreement; a rule returning “OK to use” to every TACTiC participant would have achieved at least 80.3% (≥960 of 1,196) [7, 22]. The two values are not comparable with one another. ^b^ With the prespecified mitigation plan applied; 96.0% (93.9–97.5) without [7]. ^c^ First co-primary endpoint (Overall Correct Initial Technology-Assisted Self-Selection (TASS) Outcome), with a priori and post-study mitigations applied as defined in the statistical analysis plan (Section 3.2.3.1, pp. 34–36; mitigated terms MR_AP_ and MR_PS_ in the endpoint formula at Section 4.2.2, p. 51); 84.2% (82.0–86.2) without, derived by the author. The second co-primary endpoint assesses correct use over six months rather than self-selection and is not included here [24]. ^d^ Reported in a post hoc analysis of the 431 participants without a history of cardiovascular disease (the primary prevention group) as 88.0% correct selectors and 96.2% correct rejecters, rather than as prespecified endpoints or in accuracy nomenclature; given in the Results text (p. 1120) and not in a table. The cross-tabulation from which the author’s values are derived is Table 6, p. 1120 [7]. ᵉ Derived by the author from the published cross-tabulation. Intervals are Clopper-Pearson exact. ^f^ Not reported as a single specificity estimate. The briefing materials report, for each of the seventeen label ineligibility criteria, the number and proportion of consumers meeting that criterion who nonetheless self-assessed the product as appropriate for their use (Table 11, p. 44, LDL-C arm; Table 12, pp. 45–46, total cholesterol arm) [27]; correct rejection within each warned-against subgroup is the complement of the figure shown. ^g^ In TACTiC, consumers receiving a “Do Not Use” outcome were classified as pre-screen failures and did not receive a clinician assessment. Cells C and D are therefore unpopulated and these measures cannot be estimated at any sample size.

| Measure | Accuracy term | Listed as illustrative endpoint | Reported by SELECT | Reported by CREST | Reported by TACTiC | CREST value, N = 500 (95% CI) | TACTiC value, N = 1,196 (95% CI) |
| --- | --- | --- | --- | --- | --- | --- | --- |
| Overall correct self-selection | Overall agreement<sup>a</sup> | Yes | No | Yes | Yes | 96.2% (94.1–97.7)<sup>b</sup> | 90.7% (88.9–92.3)<sup>c</sup> |
| Correct selection among those selecting to take the drug | Positive predictive value | Yes | Yes | Yes<sup>d</sup> | No | 88.5% (69.8–97.6)<sup>c</sup> | Not estimable<sup>g</sup> |
| Correct selection among those who should not take the drug | Specificity | Yes | Yes<sup>f</sup> | Yes<sup>d</sup> | No | 99.35% (98.1–99.9)<sup>c</sup> | Not estimable<sup>g</sup> |
| Correct selection among those who should take the drug | Sensitivity | No | No | No | No | 59.0% (42.1–74.4)<sup>c</sup> | Not estimable<sup>g</sup> |
| Clinician-eligible proportion (prevalence) | Prevalence | No | — | — | — | 7.8% (5.6–10.5)<sup>c</sup> | ≥80.3%<sup>a</sup> |

**Table 2.** Technology-assisted self-selection outcomes in the CREST completers population (N = 500): published cross-tabulation of participant against clinician assessment, and derived accuracy measures [7]. Derived intervals are Clopper-Pearson exact. ^a^ Denominator restricted to the 37 clinician-eligible participants receiving one of the two direct outcomes. ^b^ Denominator is the 472 participants self-selecting “not right for you”; the two participants self-selecting “ask a doctor” are excluded. The two rows differ by the single clinician “ask a doctor” participant, reclassified as a correct rejection under the prespecified mitigation plan (Section 2.3). ^c^ A rule returning “not right for you” to every consumer.

| **Clinician-verified outcome** | **Participant "OK to use"** | **Participant "Ask a doctor"** | **Participant "Not right for you"** | **Total** |
| --- | --- | --- | --- | --- |
| "OK to use" | 23 (88.5) | 2 (100) | 14 (3.0) | 39 |
| "Ask a doctor" | 0 | 0 | 1 (0.2) | 1 |
| "Not right for you" | 3 (11.5) | 0 | 457 (96.8) | 460 |
| **Total** | **26** | **2** | **472** | **500** |

Values are n (column %). Outcomes are shown before application of the prespecified mitigation plan.

*Panel B. Reported and derived measures*
| **Measure** | **Value (95% CI)** | **Source** |
| --- | --- | --- |
| Overall agreement, with mitigation | 96.2% (94.1–97.7) | Reported |
| Overall agreement, without mitigation | 96.0% (93.9–97.5) | Reported |
| Weighted kappa | 0.70 (0.58–0.83) | Reported |
| Prevalence of clinician-determined eligibility | 7.8% (5.6–10.5) | Derived |
| Sensitivity | 59.0% (42.1–74.4) | Derived |
| Sensitivity, excluding "ask a doctor" outcomes<sup>a</sup> | 62.2% (44.8–77.5) | Derived |
| Specificity | 99.35% (98.1–99.9) | Derived |
| Positive predictive value | 88.5% (69.8–97.6) | Derived |
| Negative predictive value, before mitigation<sup>b</sup> | 96.8% (94.8–98.2) | Derived |
| Negative predictive value, with prespecified mitigation<sup>b</sup> | 97.0% (95.1–98.4) | Derived |
| Agreement achieved by a null classifier<sup>c</sup> | 92.0% (89.3–94.2) | Derived |

### 2.6 Reporting and software

No reporting guideline in the EQUATOR network governs secondary analysis of published trial data combined with regulatory document review. We therefore report against no single standard, but adopted the reporting elements of STARD [33] applicable to the accuracy component, specifically the specification of index test, reference standard, participant flow, and reporting of accuracy estimates with confidence intervals.

All calculations were performed in Python 3.12.3 (Python Software Foundation) with NumPy 2.4.4 and SciPy 1.17.1; figures were produced with Matplotlib 3.10.8. Analysis code is available at https://github.com/kmolinari-source/doa-analysis-py, archived at Zenodo (https://doi.org/10.5281/zenodo.22291976).

## 3. Results

### 3.1 The endpoint options in FDA guidance do not include the false-negative rate

Section III.C.1 of the 2013 self-selection guidance [23] presents the two-by-two table of self-selection outcomes described in Section 2.2 (Figure 1), labeling cells A and D correct and cells B and C incorrect [23].

It offers three illustrative primary endpoints: overall correct self-selection, (A+D)/(A+B+C+D); correct selection among those who select to take the drug, A/(A+B); and correct self-selection among those who cannot take it, D/(B+D) [23], which are overall agreement, positive predictive value, and specificity.

The fourth quantity that the table permits, A/(A+C), does not appear [23]. This is sensitivity: the proportion of participants for whom the product is appropriate who correctly identify themselves as such. It is the only one of the four measures that could quantify how often an eligible consumer is denied access.

The set is illustrative rather than exhaustive: the guidance states that several analyses may be of interest, that one or two are usually chosen as primary endpoints, and that the choice depends on the study design and on the issues of greatest concern for nonprescription use [23].

The three studies examined here each report from the illustrative set [7, 13, 22]. SELECT reported accuracy among consumers who self-assessed the drug as appropriate for their use, A/(A+B), and, for each labeled ineligibility criterion, the proportion of consumers meeting it who nonetheless selected the product. CREST reported overall concordance, (A+D)/N, and post hoc correct selectors and correct rejecters. TACTiC reported overall correct self-selection. No OTC statin program has yet reported A/(A+C), despite the 52% to 72% range of correct self selection across the studies [7, 10–12].

The FDA’s operative framework carries the same orientation forward. The final rule defines an additional condition by its purpose, “to ensure appropriate self-selection or appropriate actual use, or both,” and requires demonstration that consumers appropriately self-select or use the product with the condition in place [18]. We did not identify, in the rule or in the procedures manual governing prescription-to-nonprescription switches, a provision addressing the rate at which such a condition denies access to consumers for whom the product is indicated [18, 21].

### 3.2 CREST: high aggregate concordance, low sensitivity, and positive net benefit

CREST reports a complete cross-tabulation of participant against clinician outcomes for all 500 completers [7]. Clinicians deemed 39 participants (7.8%) appropriate for treatment, 460 (92.0%) inappropriate, and 1 requiring physician consultation [7].

Reported concordance was 481 of 500 (96.2%; 95% CI 94.1 to 97.7) with the prespecified mitigation plan applied, and 480 of 500 (96.0%; 95% CI 93.9 to 97.5) without [7]. The investigators additionally reported a weighted kappa of 0.70 (95% CI 0.58 to 0.83), which corrects for chance agreement [7].

Derived from the cross-tabulation, specificity was 458 of 461 (99.35%; 95% CI 98.11 to 99.87).Sensitivity was 23 of 39 (59.0%; 95% CI 42.1 to 74.4); restricting the denominator to participants receiving one of the two direct outcomes, 23 of 37 (62.2%). Positive predictive value was 23 of 26 (88.5%) and negative predictive value 457 of 472 (96.8%). Fourteen participants whom clinicians deemed appropriate for treatment received a “not right for you” outcome, and two received “ask a doctor.” [7]

The reported positive predictive value and the derived sensitivity differ by 29.5 percentage points (95% CI 11.8 to 47.4) and are calculated from the same 500 participants [7]. The published post hoc analysis in the 431 primary prevention participants reports the former, at 88.0% correct selectors, and does not report the latter [7].

Overall agreement in a self-selection study depends on the base rate of eligibility in the enrolled population. Because 92.0% of CREST participants were clinician-ineligible, a null classifier returning “not right for you” to every consumer would have achieved 92.0% agreement; the reported 96.2% exceeds that baseline by 4.2 percentage points [7]. The corresponding baseline runs in the opposite direction in TACTiC, where enrollment was restricted to consumers the application had approved: a null classifier returning “OK to use” to every participant would have achieved at least 80.3%, against a reported 90.7% [22]. The two reported figures are therefore not comparable with one another, and neither is interpretable without its base rate.

Decision curve analysis nonetheless favors the application: across the reasonable range of 5% to 20% its net benefit exceeded that of both comparators at every threshold (Figure 2), and both crossovers lie outside the range, at 3.4% against unscreened access and 88.5% against no access [7]. Only below approximately 3.4%, where inappropriate exposure is valued as nearly harmless, does unscreened access produce greater net benefit. No decision curve can be constructed for TACTiC: the net benefit of unscreened access depends on the number of eligible consumers among the rejected, which the design left unmeasured.

**Figure 2.**
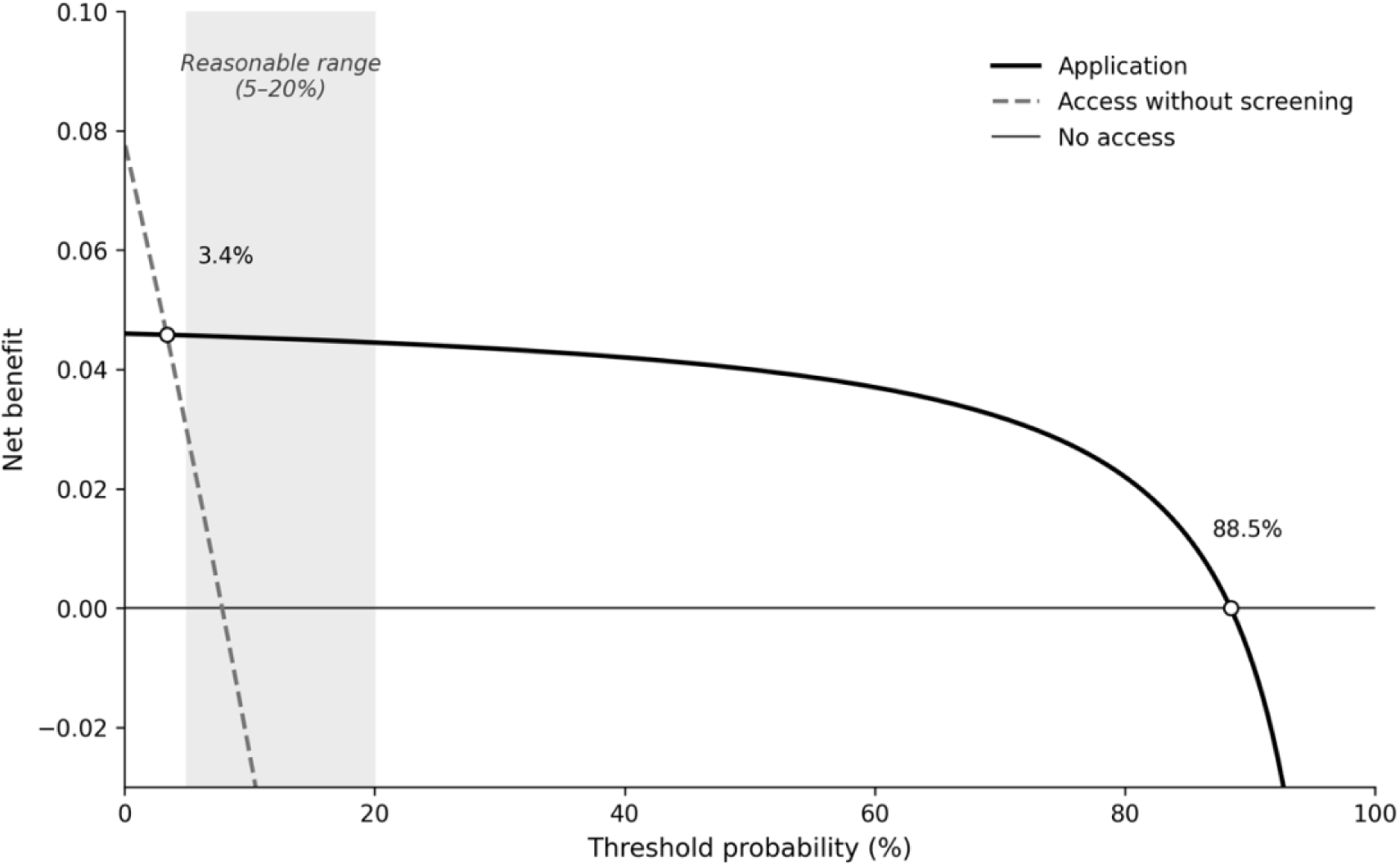
Decision curves for the CREST application against two comparator strategies. Net benefit of the application (solid line), of granting access without screening (dashed line), and of granting no access (horizontal line at zero), across threshold probabilities from 0% to 100%, calculated from the published cross-tabulation (Table 2, Panel A). The shaded band marks the reasonable range of 5% to 20% to which primary interpretation is restricted. The application curve crosses the unscreened access curve at 3.4% and the no access line at 88.5%; both crossings lie outside the band. The vertical axis is truncated at −0.03; the unscreened access curve continues below the plotted range.

### 3.3 TACTiC cannot estimate the false-negative rate by construction

Of 12,624 consumers who completed the web application assessment in TACTiC, 10,332 received a “Do Not Use” outcome [22]; none was enrolled or received a clinician assessment [8, 22]. The statistical analysis plan classifies them as pre-screen failures (statistical analysis plan, Section 3.1, p. 25; clinical study protocol, Section 9.3, pp. 68–69), one of nine categories of consumer who did not sign the consent form (Figure 3) [8, 24], including consumers the application approved who chose not to participate; what distinguishes the rejected is not the label but that no reference standard was applied to any of them (Figure 4).

**Figure 3.**
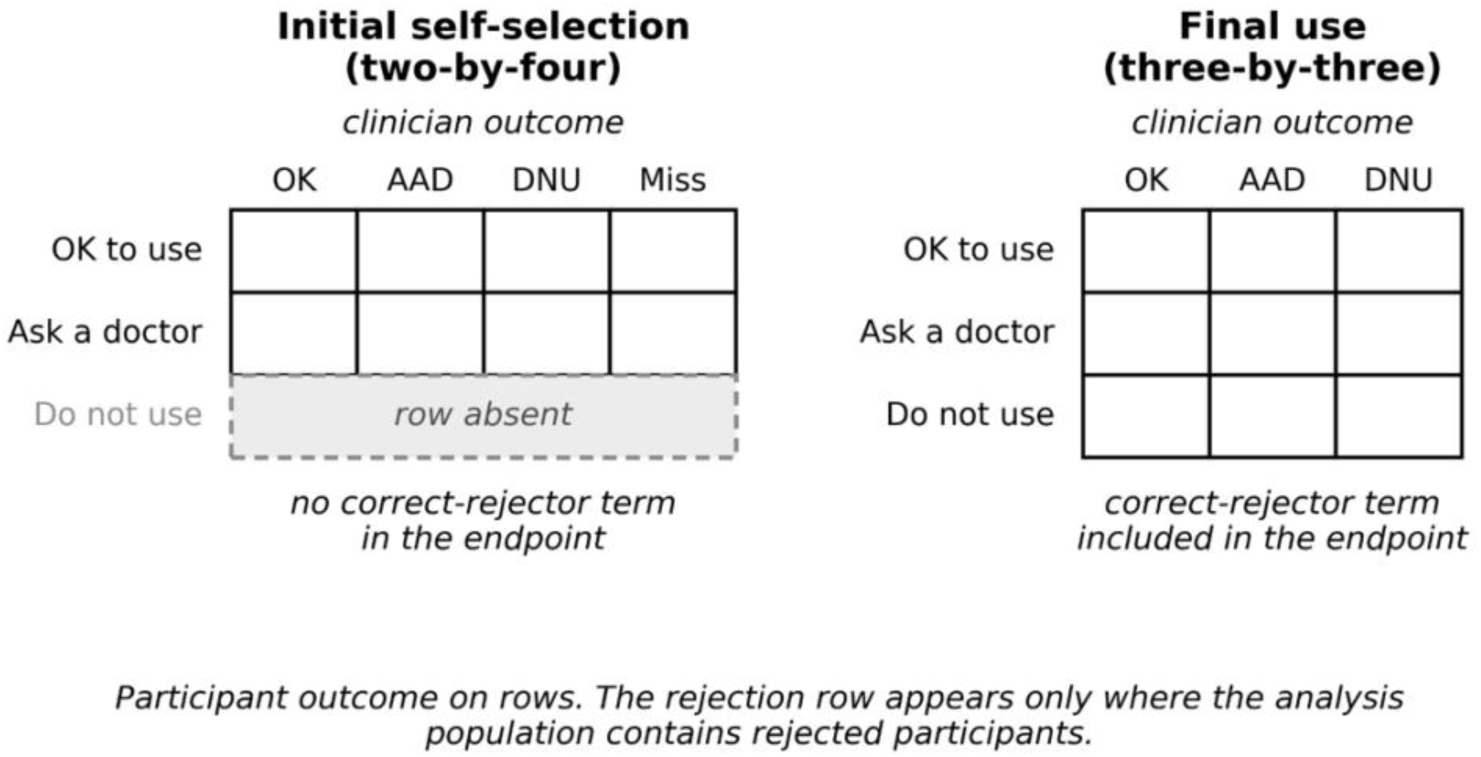
Outcome tables illustrating possible self-selection outcomes in the TACTiC statistical analysis plan, for the initial self-selection endpoint (Section 4.2.2, Figure 1, p. 51) and the final use endpoint (Section 4.2.2, Figure 2, p. 53) [24]. Participant outcome is shown on rows and clinician outcome on columns. The initial self-selection table contains no participant rejection row, and the corresponding endpoint numerator contains no correct-rejector term. OK, OK to use; AAD, ask a doctor; DNU, do not use; Miss, missing.

**Figure 4.**
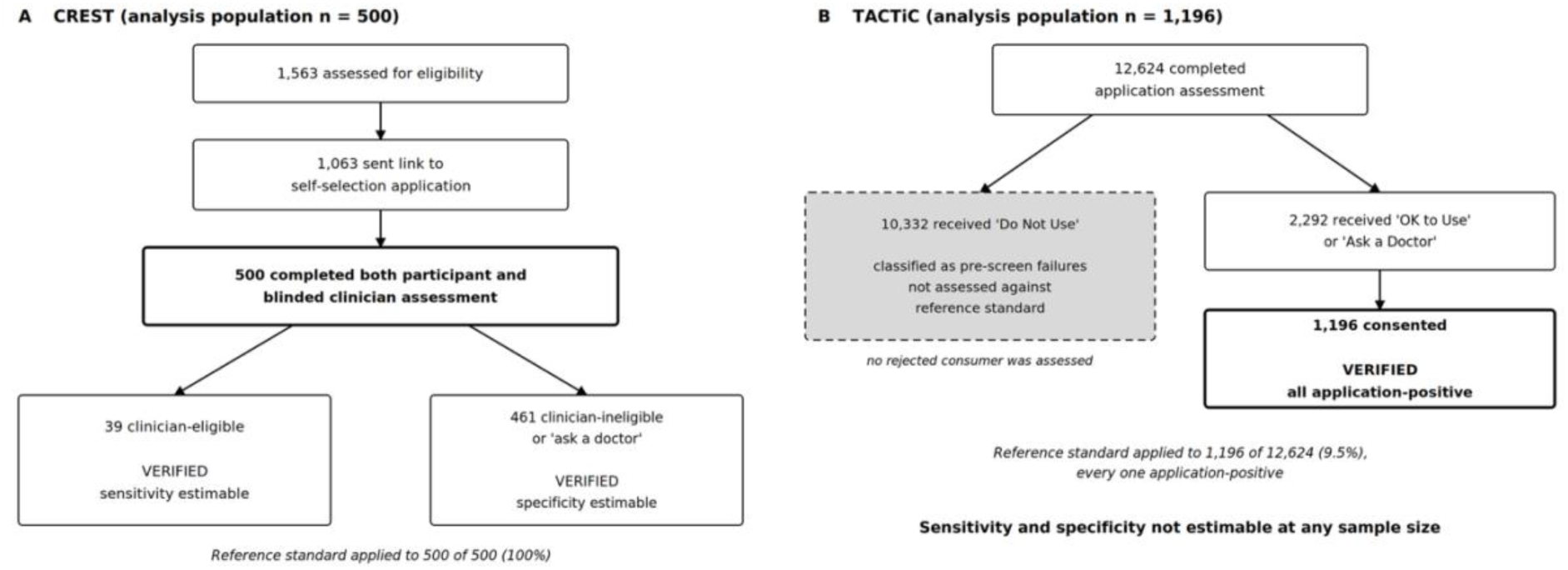
Application of the reference standard in CREST and TACTiC. **(A)** In CREST, all 500 participants who completed the study received both a technology-assisted self-assessment and an independent assessment by a clinician blinded to the participant’s entries. Both clinician-eligible and clinician-ineligible participants were verified, and sensitivity and specificity are therefore both estimable. The right-hand box combines the 460 participants whom clinicians deemed inappropriate for treatment with the single participant assigned an “ask a doctor” outcome. **(B)** In TACTiC, consumers receiving a “Do Not Use” outcome from the application were classified as pre-screen failures (statistical analysis plan, Section 3.1, p. 25) [24], were not enrolled, and received no clinician assessment; the shaded box with a dashed border denotes this group. Every participant who received the reference standard had already been approved by the application, so no cell of the cross-tabulation corresponding to a rejected consumer is populated in the initial self-selection analysis. Counts are as reported in the participant flow diagrams of the two publications [7, 22].

The remaining 2,292 received “OK to Use” or “Ask a Doctor,” of whom 1,196 (52.2%) signed informed consent [22]. The reference standard was therefore applied to 1,196 of 12,624 application completers (9.5%), every one of them application-positive [22].

The endpoint architecture reflects this directly (Figure 3): the statistical analysis plan’s illustration of initial self-selection outcomes has no participant “Do Not Use” row and the first co-primary numerator no term for correct rejection, whereas the final use illustration and the second co-primary numerator include both (Section 4.2.2, Figures 1 and 2, pp. 51 and 53; Supplementary Methods S6) [24]. Correct rejection is measured where the analysis population permits it and absent where it does not.

No participant whom the application turned away was assessed against the reference standard [22]. Neither sensitivity nor specificity can be estimated from TACTiC at any sample size, and the study contributes no information about the rate at which eligible consumers are denied access.

### 3.4 Both of TACTiC’s co-primary endpoints depend on mitigation; second-endpoint mitigations are discontinuation

The TACTiC primary endpoints are defined to include mitigated outcomes as correct [8, 24]. The success criterion, prespecified in the statistical analysis plan, required the lower bound of a two-sided 95% Clopper-Pearson interval to exceed 85% [8, 24].

Reported results met that criterion. The first co-primary was 1,085 of 1,196 (90.7%; 95% CI 88.9 to 92.3) and the second 1,132 of 1,154 (98.1%; 95% CI 97.1 to 98.8) [22, 25]. Our recomputation reproduces both published intervals exactly.

Excluding mitigated participants, the first co-primary is 1,007 of 1,196 (84.2%; 95% CI 82.0 to 86.2) and the second 934 of 1,154 (80.9%; 95% CI 78.5 to 83.2) [22]. Neither lower bound exceeds 85%. Of the 78 mitigations applied to the first endpoint, 34 were required to bring the lower bound above the threshold; of the 198 applied to the second, 71. The composition of the second endpoint’s mitigations is reported in the publication’s supplemental material (Supplemental Table 2) [22]. Sixty participants (5.2% of the analysis population) stopped use despite being eligible to continue treatment. A further 73 (6.3%) stopped for reasons not confirmed by a clinician, of whom 54 reported muscle symptoms and 12 liver symptoms. Ninety (7.8%) were mitigated for correct use with a missing LDL-C retest. Categories are not mutually exclusive [22]. Participants who discontinued therapy therefore account for 133 of the 198 mitigations counted toward a reported 98.1% correct use rate [22].

Appendix D of the statistical analysis plan (Use and Re-Selection Mitigations, pp. 92–93), paralleled in the clinical study protocol (Table 6, pp. 74–75), states the rationale for the two a priori mitigations in parallel terms, treating an incorrect affirmative response and a decision to stop treatment alike as conservative outcomes from which no medical harm can come to the participant (quoted in this paper’s Supplementary Methods S5) [8, 24].

By contrast, CREST reported both of its endpoints with and without mitigation, in its primary and secondary endpoint tables [7]. The TACTiC statistical analysis plan prespecified the unmitigated analysis twice, in the primary analysis of the first co-primary endpoint and again under a sensitivity analysis, and specified a table of mitigation counts by descriptor distinguishing a priori from post-study mitigations across three analysis populations (Section 4.2.2, pp. 52 and 55; passages quoted in this paper’s Supplementary Methods S4) [24]. The published supplemental material reports descriptors for a single population without that distinction [22], and we did not identify the unmitigated analyses in the publication, its supplemental material, the protocol, the statistical analysis plan, or the ClinicalTrials.gov results record [8, 22, 24, 25].

## 4. Discussion

### 4.1 Principal findings

The 2013 FDA guidance on self-selection studies presents a two-by-two table of consumer decisions and offers three illustrative primary endpoints, none of which is the proportion of eligible consumers who correctly identify themselves as such [23]. All three published consumer self-selection studies of nonprescription statins report from that illustrative set, and none reports that proportion, though CREST published the table from which it can be recovered [7, 13, 22]. Recovered from that table, sensitivity was 59.0% against a specificity of 99.35%, and the reported positive predictive value of 88.5% exceeds the derived sensitivity by 29.5 percentage points in the same 500 participants. In TACTiC, consumers whom the application turned away were classified as pre-screen failures and never assessed against the reference standard, so the quantity cannot be recovered at any sample size. Both TACTiC co-primary endpoints fall below their prespecified success criterion when mitigated participants are excluded, and 133 of the 198 mitigations applied to the second endpoint are participants who stopped taking the drug. The population definitions governing the reported persistence figure exclude, by construction, anyone who discontinued.

### 4.2 Where the asymmetry originates

None of this departs from accepted practice. The TACTiC mitigation framework was prespecified in the protocol and statistical analysis plan, built into the endpoint definition rather than applied afterward, coded by an independent contract research organization, adjudicated by dual coding with third-party resolution of disagreements, and conducted with the medical teams blinded to the effect of post-study mitigations on the result [8, 22, 24]. The statistical analysis plan cites the 2013 guidance directly [23, 24]. CREST likewise prespecified its mitigation plan, reported both endpoints with and without it, and stated its principal limitation in its own discussion [7]. The sponsors did what the guidance asks.

The asymmetry sits upstream, in what the evaluative framework counts as an error. A consumer who takes a product inappropriately is an identifiable person exposed to an identifiable hazard, and the endpoints, the study designs, and the advisory committee record all address that outcome directly: in 2005 the committee voted unanimously that user behavior after initiation raised concerns about safe and effective nonprescription use [12], and in TACTiC discontinuation while eligible is counted toward the endpoint measuring that behavior [22, 24]. A consumer who is incorrectly turned away is not enrolled, not counted, and not followed. The statistical analysis plan states the premise without ambiguity, applying identical reasoning to a response that disqualifies and to an action that discontinues: in each case the conservative outcome means no medical harm can come to the participant [24]. Its own name for the outcome in which a participant stops and discontinues from the study while the clinician judged them “OK to Use” is “Lost Opportunity” (Appendix D, p. 93; clinical study protocol, Table 6, p. 75), and it classifies that outcome as correct [8, 24]. Under a framework oriented to preventing inappropriate use, that reasoning is sound: the nonprescription approval standard asks whether a product can be used safely and effectively without professional supervision, and an agency applying it answers for exposures it permits, not for treatment a consumer forgoes. The asymmetry is therefore not an oversight but a consequence of the question the framework was built to answer. It becomes a problem only when the same evidence is offered for a distinct claim, that nonprescription availability will reduce undertreatment, which the framework does not test; under the public health rationale offered for the switch, that is the outcome of interest.

### 4.3 Why the false negative is not costless

The standard reply is that a consumer denied access can consult a physician. That reply assumes the availability of the resource whose absence justifies the switch: the rule exists to increase consumer access [18], the trial literature attributes undertreatment in part to limited engagement with routine care [22], and PALM registry data place 36.6% of statin-eligible primary prevention patients on no statin, the complement of the 63.4% treated, even within practices where they are established patients [3]. For a consumer in that position an erroneous rejection is not a referral but an exit; in CREST such errors befell 41% of clinician-eligible participants, the complement of its sensitivity [7].

The discontinuation data make the same point without the inference. Fifty-four participants stopped therapy for muscle symptoms a clinician did not confirm, and twelve for unconfirmed liver symptoms [22]; randomized crossover trials in which each participant alternated blinded statin and placebo periods indicate that most symptoms attributed to statins occur equally on placebo [35, 36], and TACTiC’s single-arm open-label design cannot distinguish drug effect from nocebo; neither can the application, which received only the symptom as reported. Each left a therapy for which they remained eligible on the basis of unverified symptoms and was counted toward a reported correct use rate of 98.1% [22]. Stopping is not clinically neutral: statin nonadherence is associated with diminished LDL-C lowering and with adverse cardiovascular outcomes [5].

The clinical standard itself is also moving. TACTiC’s application encodes the 2018 American College of Cardiology/American Heart Association (ACC/AHA) cholesterol guideline’s pooled cohort equations [22]; the 2026 guideline replaced those equations with the PREVENT-ASCVD models and revised the risk thresholds that govern statin initiation [2], so the eligibility the application enforces and the eligibility current guidelines define have already diverged. A national survey analysis estimates that the 2026 ACC/AHA multisociety dyslipidemia guideline expands statin eligibility by an estimated 24.5 million adults relative to 2018, largely through 30-year risk assessment the application does not perform [37], so the divergence runs toward denial; its size within the application’s own population is unmeasured, as is every other component of the false-negative rate.

### 4.4 Implications for applications under the additional conditions framework

The additional conditions for nonprescription use pathway permits a sponsor to place a technological gate between the consumer and the product [18] but does not specify what the gate must be shown to do. If the evidentiary expectations carried forward are those illustrated in the 2013 guidance [23], an application can satisfy them without ever estimating how often an eligible consumer is refused; the final rule does not displace those expectations, and its demonstration requirement runs in the same direction [18].

Four changes to how self-selection studies are designed and reported would close that gap, the absence of any measurement of denial. None requires a new methodology, and three of the four were already planned within the programs examined here.

First, apply the reference standard to a sample of consumers the tool rejects. A verification subsample of application-negatives, assessed by the same blinded clinician procedure as application-positives, would permit estimation of sensitivity and specificity. This is the one change that requires a design decision rather than a reporting one, and it is the change CREST’s investigators themselves called for in recommending studies that enrich for clinician-confirmed eligible participants, so that the application’s performance among them can be estimated [7]. TACTiC instead selected its population by the application’s own approvals, making such estimation structurally impossible: its analysis population contains no rejected participant, and its endpoint illustration accordingly has no rejection row [24]. Feasibility of reaching rejected consumers is not the obstacle: the TACTiC protocol specified a telephone sub-study of untreated pre-screen failures at high cardiovascular risk, the very consumers the application had turned away, to survey their treatment-seeking behavior (Section 9.4.4 and Appendix H) [8]. The infrastructure to reach rejected consumers existed; it was not used to apply the reference standard.

Second, report sensitivity alongside overall correct selection wherever the design yields a cross-tabulation, as CREST’s did. Both are computable from the same table, and in CREST reporting one without the other concealed a difference of nearly thirty percentage points [7]. For designs like TACTiC’s, this reporting becomes possible only once the first change is made. Third, report co-primary endpoints with and without mitigations. This is not a new demand: CREST published both endpoints both ways, and TACTiC’s own statistical analysis plan prespecified the unmitigated analysis twice and a mitigation-descriptor table across three populations (Section 3.4) [7, 24]. The plan states the intent directly: “The impact of the a priori and post-study mitigations on the overall co-primary endpoints will be assessed by evaluating the endpoints with and without these mitigations” (Section 3.2.3.1, p. 35) [24]. We did not identify these analyses in any public document. The request is therefore not that sponsors generate new evidence, but that they publish analyses their own plans already specify in detail. Publication matters because the unmitigated figures are the only direct measure of how much of each endpoint the mitigation layer supplies, 6.5 and 17.2 percentage points across the two endpoints (Table 3); without them that contribution is invisible to the committees and consumers weighing the evidence.

**Table 3.** TACTiC co-primary endpoints against the prespecified success criterion, with and without mitigations [22]. Success criterion, prespecified in the clinical study protocol (Section 9.1, p. 65) and statistical analysis plan (Section 1.3, pp. 21–22): H₀ P ≤ 85% versus H₁ P > 85%, rejected only if the lower bound of the two-sided 95% Clopper-Pearson interval exceeds 85%. ^a^ True outcome rate assumed in the sample size justification (statistical analysis plan, Section 1.3, pp. 21– 22). ^b^ Derived by the author by removing mitigated participants from each endpoint’s numerator of correct outcomes, with mitigation counts as reported in the publication’s supplemental tables [22]. Of the 78 mitigations applied to the first endpoint, 34 were required for the lower bound to exceed 85%; of the 198 applied to the second, 71. Intervals for the reported endpoints were reproduced exactly by our method. ^c^ Analysis populations are those named in the statistical analysis plan (Section 2.1, pp. 23–25) and clinical study protocol (Section 9.3, Table 4, pp. 68–69). Self-Selection (SS) population: all participants who signed the informed consent form (N = 1,196), prespecified as the primary analysis population for the first co-primary endpoint (statistical analysis plan, Section 4.2.2, p. 50; protocol, Section 9.4.2.1, p. 79). Per Protocol (PP) population: participants in the actual use study intent-to-treat population who completed Virtual Visit 2, or had sufficient medical and medication history and, where required, a verified LDL-C value, to determine a final use outcome (n = 1,154), prespecified as the primary analysis population for the second co-primary endpoint (same sections). The actual use study intent-to-treat population is a distinct and larger set (n = 1,188) and is the denominator of neither co-primary endpoint as reported.

| Endpoint | Planning assumption<sup>a</sup> | With mitigations | Criterion met | Without mitigations (derived)<sup>b</sup> | Criterion met |
| --- | --- | --- | --- | --- | --- |
| Overall Correct Initial TASS Outcome, Self-Selection (SS) population (N = 1,196)<sup>c</sup> | 88.2–90.0% | 1,085<br>(90.7%; 95% CI 88.9–92.3) | Yes | 1,007<br>(84.2%; 95% CI 82.0–86.2) | No |
| Overall Correct Final Use Outcome, Per Protocol (PP) population (n = 1,154)<sup>c</sup> | 90.0% | 1,132<br>(98.1%; 95% CI 97.1–98.8) | Yes | 934<br>(80.9%; 95% CI 78.5–83.2) | No |

Success criterion, prespecified in the clinical study protocol (Section 9.1, p. 65) and statistical analysis plan (Section 1.3, pp. 21–22): $H_0 P \leq 85\%$ versus $H_1 P > 85\%$ , rejected only if the lower bound of the two-sided 95% Clopper-Pearson interval exceeds 85%.

**Table 4.**
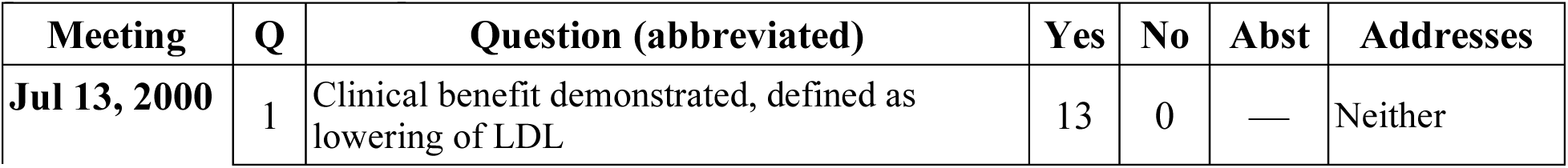
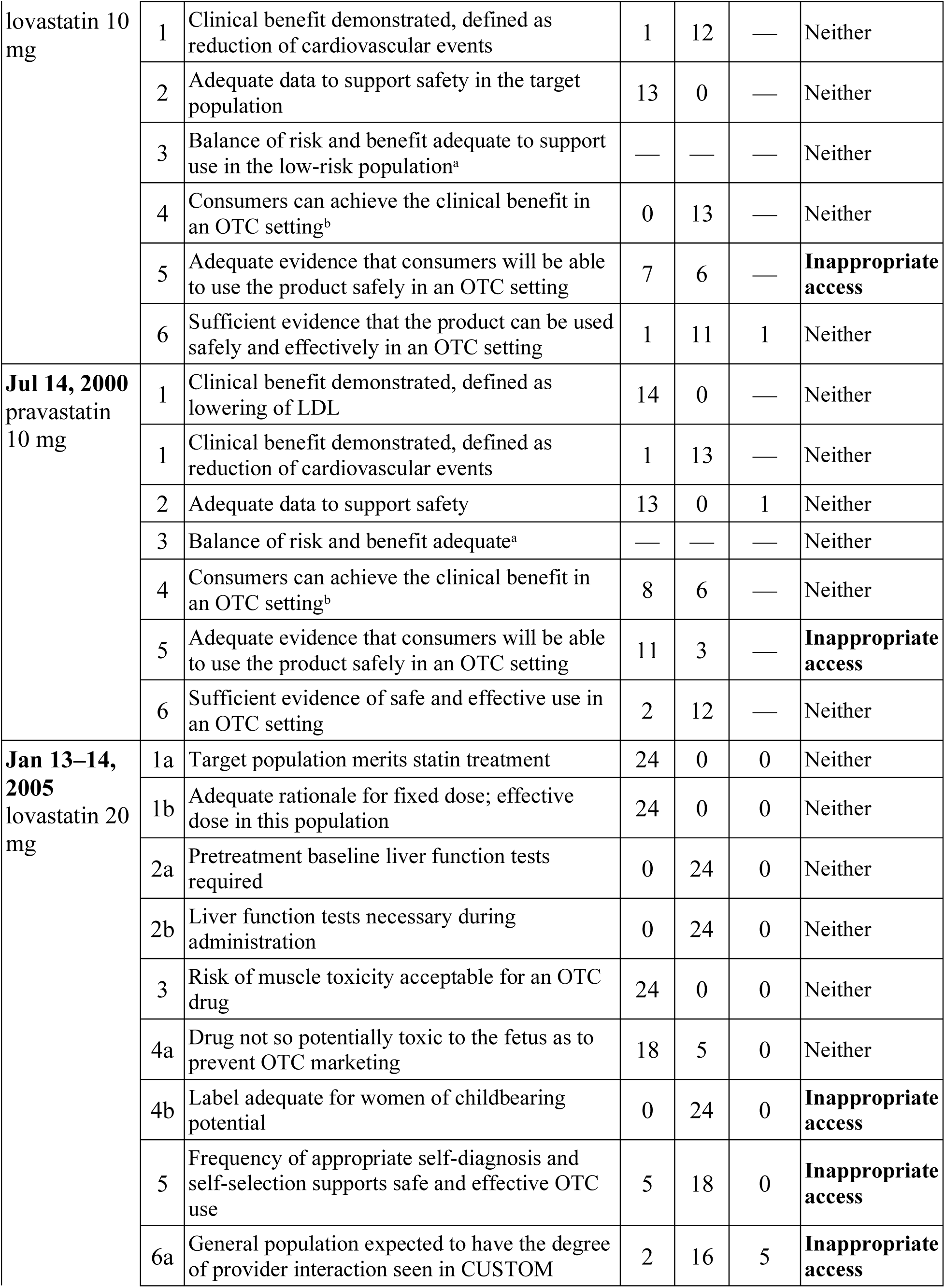
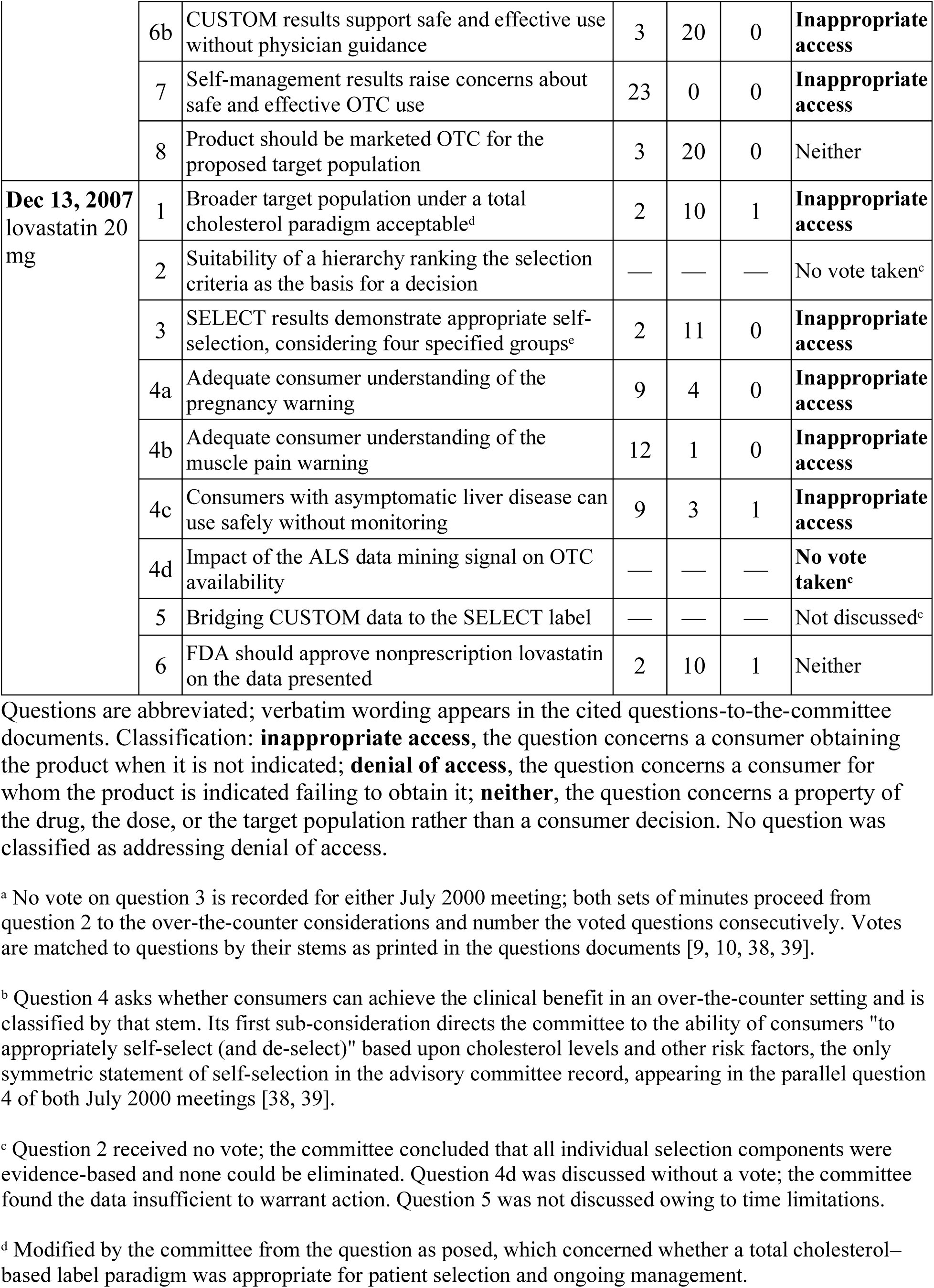
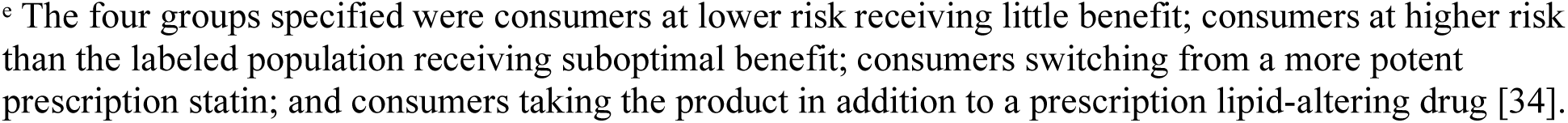
Questions put to joint advisory committees for the four nonprescription statin applications, with committee votes and classification of each question by the error it addresses [9, 10, 12, 14, 34, 38, 39]. Questions are abbreviated; verbatim wording appears in the cited questions-to-the-committee documents. Classification: **inappropriate access**, the question concerns a consumer obtaining the product when it is not indicated; **denial of access**, the question concerns a consumer for whom the product is indicated failing to obtain it; **neither**, the question concerns a property of the drug, the dose, or the target population rather than a consumer decision. No question was classified as addressing denial of access. ^a^ No vote on question 3 is recorded for either July 2000 meeting; both sets of minutes proceed from question 2 to the over-the-counter considerations and number the voted questions consecutively. Votes are matched to questions by their stems as printed in the questions documents [9, 10, 38, 39]. ^b^ Question 4 asks whether consumers can achieve the clinical benefit in an over-the-counter setting and is classified by that stem. Its first sub-consideration directs the committee to the ability of consumers “to appropriately self-select (and de-select)” based upon cholesterol levels and other risk factors, the only symmetric statement of self-selection in the advisory committee record, appearing in the parallel question 4 of both July 2000 meetings [38, 39]. ^c^ Question 2 received no vote; the committee concluded that all individual selection components were evidence-based and none could be eliminated. Question 4d was discussed without a vote; the committee found the data insufficient to warrant action. Question 5 was not discussed owing to time limitations. ^d^ Modified by the committee from the question as posed, which concerned whether a total cholesterol– based label paradigm was appropriate for patient selection and ongoing management. ^e^ The four groups specified were consumers at lower risk receiving little benefit; consumers at higher risk than the labeled population receiving suboptimal benefit; consumers switching from a more potent prescription statin; and consumers taking the product in addition to a prescription lipid-altering drug [34].

| Meeting | Q | Question (abbreviated) | Yes | No | Abst | Addresses |
| --- | --- | --- | --- | --- | --- | --- |
| Jul 13, 2000 | 1 | Clinical benefit demonstrated, defined as lowering of LDL | 13 | 0 | — | Neither |
| lovastatin 10 mg | 1 | Clinical benefit demonstrated, defined as reduction of cardiovascular events | 1 | 12 | — | Neither |
|  | 2 | Adequate data to support safety in the target population | 13 | 0 | — | Neither |
|  | 3 | Balance of risk and benefit adequate to support use in the low-risk population<sup>a</sup> | — | — | — | Neither |
|  | 4 | Consumers can achieve the clinical benefit in an OTC setting<sup>b</sup> | 0 | 13 | — | Neither |
|  | 5 | Adequate evidence that consumers will be able to use the product safely in an OTC setting | 7 | 6 | — | **Inappropriate access** |
|  | 6 | Sufficient evidence that the product can be used safely and effectively in an OTC setting | 1 | 11 | 1 | Neither |
| **Jul 14, 2000**<br>pravastatin 10 mg | 1 | Clinical benefit demonstrated, defined as lowering of LDL | 14 | 0 | — | Neither |
|  | 1 | Clinical benefit demonstrated, defined as reduction of cardiovascular events | 1 | 13 | — | Neither |
|  | 2 | Adequate data to support safety | 13 | 0 | 1 | Neither |
|  | 3 | Balance of risk and benefit adequate<sup>a</sup> | — | — | — | Neither |
|  | 4 | Consumers can achieve the clinical benefit in an OTC setting<sup>b</sup> | 8 | 6 | — | Neither |
|  | 5 | Adequate evidence that consumers will be able to use the product safely in an OTC setting | 11 | 3 | — | **Inappropriate access** |
|  | 6 | Sufficient evidence of safe and effective use in an OTC setting | 2 | 12 | — | Neither |
| **Jan 13–14, 2005**<br>lovastatin 20 mg | 1a | Target population merits statin treatment | 24 | 0 | 0 | Neither |
|  | 1b | Adequate rationale for fixed dose; effective dose in this population | 24 | 0 | 0 | Neither |
|  | 2a | Pretreatment baseline liver function tests required | 0 | 24 | 0 | Neither |
|  | 2b | Liver function tests necessary during administration | 0 | 24 | 0 | Neither |
|  | 3 | Risk of muscle toxicity acceptable for an OTC drug | 24 | 0 | 0 | Neither |
|  | 4a | Drug not so potentially toxic to the fetus as to prevent OTC marketing | 18 | 5 | 0 | Neither |
|  | 4b | Label adequate for women of childbearing potential | 0 | 24 | 0 | **Inappropriate access** |
|  | 5 | Frequency of appropriate self-diagnosis and self-selection supports safe and effective OTC use | 5 | 18 | 0 | **Inappropriate access** |
|  | 6a | General population expected to have the degree of provider interaction seen in CUSTOM | 2 | 16 | 5 | **Inappropriate access** |
|  | 6b | CUSTOM results support safe and effective use without physician guidance | 3 | 20 | 0 | **Inappropriate access** |
|  | 7 | Self-management results raise concerns about safe and effective OTC use | 23 | 0 | 0 | **Inappropriate access** |
|  | 8 | Product should be marketed OTC for the proposed target population | 3 | 20 | 0 | Neither |
| **Dec 13, 2007**<br>lovastatin 20 mg | 1 | Broader target population under a total cholesterol paradigm acceptable<sup>d</sup> | 2 | 10 | 1 | **Inappropriate access** |
|  | 2 | Suitability of a hierarchy ranking the selection criteria as the basis for a decision | — | — | — | No vote taken<sup>e</sup> |
|  | 3 | SELECT results demonstrate appropriate self-selection, considering four specified groups<sup>e</sup> | 2 | 11 | 0 | **Inappropriate access** |
|  | 4a | Adequate consumer understanding of the pregnancy warning | 9 | 4 | 0 | **Inappropriate access** |
|  | 4b | Adequate consumer understanding of the muscle pain warning | 12 | 1 | 0 | **Inappropriate access** |
|  | 4c | Consumers with asymptomatic liver disease can use safely without monitoring | 9 | 3 | 1 | **Inappropriate access** |
|  | 4d | Impact of the ALS data mining signal on OTC availability | — | — | — | **No vote taken<sup>e</sup>** |
|  | 5 | Bridging CUSTOM data to the SELECT label | — | — | — | Not discussed<sup>e</sup> |
|  | 6 | FDA should approve nonprescription lovastatin on the data presented | 2 | 10 | 1 | Neither |

Fourth, define mitigation categories publicly and distinguish those reflecting continued appropriate use from those reflecting discontinuation. The category holding sixty participants, “stopped use despite being eligible to continue treatment,” corresponds to the prespecified mitigation the statistical analysis plan names “Lost Opportunity” (Appendix D, p. 93; clinical study protocol, Table 6, p. 75), whose stated content is that the participant stopped and discontinued while the clinician determined they were “OK to Use.” [24] The correspondence is nowhere stated in the publication, the supplemental material uses a different label, and no source relates the category to the persistence denominator from which such participants are excluded [22]. The distinction matters because a category that merges continued appropriate use with discontinuation lets a stopped therapy count as a success; separated, the same data would show how much of correct use is actual use.

None of these would make approval harder to obtain. They would make the resulting evidence answer the question the switch is justified by.

### 4.5 Limitations

Published summary data were analyzed rather than participant-level data, and the unmitigated TACTiC endpoints are derived by subtraction; this paper’s interval method reproduces both published intervals exactly, which supports the derivation, but participant-level access would permit direct calculation.

Sensitivity in CREST rests on 39 clinician-eligible participants, and its interval is correspondingly wide (42.1% to 74.4%); the estimate is imprecise rather than a point value, and the study was neither designed nor powered for this quantity, a limitation its investigators stated [7].

Only one operating point of the CREST application was published, the sensitivity and specificity pair its deployed configuration produced. A receiver operating characteristic curve therefore cannot be constructed, and we cannot determine whether an alternative configuration would produce greater net benefit. The curves also reflect CREST’s sample prevalence of 7.8% clinician-eligible participants, itself shaped by enrollment caps on age and sex [7]; the crossover against unscreened access rises with prevalence and enters the reasonable range once the eligible fraction exceeds roughly 11%.

Decision curve analysis anchors the treat-none strategy at zero net benefit [32], so an incorrectly rejected consumer contributes neither benefit nor harm; the framework used for the analysis therefore embeds the same asymmetry the paper describes, and because untreated eligibility carries cumulative cardiovascular risk [1], the analysis understates the cost of a false negative. The TACTiC ClinicalTrials.gov results record and the publication disagree on all three label-warning compliance outcomes: the registry reports two participants with a clinician-identified stop-use warning, one concordant, where the publication reports none; 46 of 59 concordant on a do-not-use warning (78.0%) against the publication’s 47 (79.7%); and 23 of 25 on an ask-a-doctor warning (92.0%) against the publication’s 20 of 24 (83.3%). This paper did not select between them; the differences are unexplained in the public record [22, 25].

## 5. Conclusions

Across the guidance’s illustrative endpoints, the questions put to four advisory committees, and the endpoints of all three published studies, evidence for consumer self-selection of nonprescription statins has been evaluated by whether ineligible consumers are excluded. In the last sixteen years, across two sponsors and instruments ranging from a printed carton to a web application, the rate at which eligible consumers are denied access has not been reported, and in the largest and most recent study it cannot be estimated. In the United Kingdom, where nonprescription simvastatin has been available through pharmacies since 2004 [40], the corresponding rate is likewise unrecorded. The 2013 guidance directs its illustrative endpoints at inappropriate use [23], as did the questions put to four advisory committees [12, 34, 38, 39], and by that measure the self-selection evidence framework works. The public health objective offered in support of a switch is fewer eligible adults untreated. Whether technology-assisted self-selection advances that objective cannot be known from evidence that counts only the errors of admission; the four changes above would let the next application answer it.

## Data Availability

All materials analyzed in this study are publicly available. The clinical study protocol and statistical analysis plan for NCT04964544 were obtained through AstraZeneca's clinical trial disclosure process. Trial results were obtained from ClinicalTrials.gov (NCT04964544, NCT01964326). FDA guidance documents, Federal Register notices, and advisory committee minutes, questions, and briefing materials were obtained from FDA.gov and the FDA Web Archive. Peer-reviewed publications are cited in full. All derived calculations, including confidence intervals and decision curve analyses, are reproducible from the values reported in the cited sources; the analysis code is archived at Zenodo (https://doi.org/10.5281/zenodo.22291976).

https://doi.org/10.1016/j.jacc.2024.03.388

https://doi.org/10.1016/j.jacc.2021.06.048

https://clinicaltrials.gov/study/NCT04964544

https://clinicaltrials.gov/study/NCT01964326

https://doi.org/10.5281/zenodo.22291976

## Ethics approval

Not applicable. This secondary analysis used only published results and publicly available regulatory documents, involved no interaction with human participants and no identifiable data, and did not require institutional review board approval. The design of each underlying trial was reviewed by the US Food and Drug Administration, and Advarra served as central institutional review board for both [7, 22].

## Supplementary information

Supplementary Methods S1 to S4, comprising the CREST handling rules and interval methods, the TACTiC interval validation, the decision curve specification, and the statistical analysis plan passages quoted, accompany the online version of this article.

## Use of AI-assisted technologies

The author used a large language model to assist with drafting and editing manuscript text. The author verified all source material, performed all data extraction and statistical calculations, and reviewed and revised all AI-assisted text. The author takes full responsibility for the accuracy, integrity, and originality of the work.

## Author contributions

Kyle Molinari is the sole author and was responsible for study conception, source identification and extraction, statistical analysis, drafting, and final approval of the manuscript, and agrees to be accountable for all aspects of the work.

## Ethics approval

Not applicable. This study analyzed publicly available published data, regulatory documents, and sponsor study documents. No human participants were involved and no institutional review board approval was required.

