## Supplementary Methods S1-S6 for "Denial of Access as an Unmeasured Outcome in Nonprescription Statin Self-Selection Studies"

Reference numbers correspond to the reference list of the main text.

### Supplementary Methods S1. Handling of the "ask a doctor" outcome, accuracy measures, and the mitigation layer in the CREST reanalysis

CREST reports a three-by-three cross-tabulation of participant against clinician outcomes for all 500 completers; Table 2 Panel A of the main text presents it with the clinician’s independent assessment as rows and the participant outcome ("OK to use," "ask a doctor," "not right for you") as columns [7]. Because the application returns three possible outcomes, reducing the published table to the guidance’s two-by-two form requires a rule for the "ask a doctor" outcome. We defined the index test as positive when the participant self-selected "OK to use," the only outcome granting access to the product, and the reference standard as positive when the clinician’s independent assessment returned "OK to use." In the primary handling rule, clinician-eligible participants who received "ask a doctor" (n = 2) were retained in the denominator of clinician-eligible participants and counted as not granted access, because that outcome does not place the product in the consumer’s hands; sensitivity was therefore 23 of 39. In the secondary rule, restricted to the two direct outcomes, these participants were excluded from the denominator; sensitivity was 23 of 37. Both are reported in Section 3.2 of the main text.

The measures were defined as follows. Sensitivity is the proportion of clinician-eligible participants who self-selected "OK to use." Specificity is the proportion of clinician-ineligible participants (the 460 assessed "not right for you" together with the single participant assessed "ask a doctor") who did not self-select "OK to use." Positive predictive value is the proportion of participants who self-selected "OK to use" whom the clinician deemed eligible (23 of 26), and negative predictive value the proportion of participants who self-selected "not right for you" whom the clinician deemed ineligible (457 of 472). Overall agreement is the proportion of participants whose outcome matched the clinician’s. The null classifier is a rule returning "not right for you" to every participant; its agreement equals the proportion of participants the clinician assessed "not right for you" (460 of 500, 92.0%).

The cross-tabulation reports outcomes before application of the prespecified mitigation plan; the primary endpoint table reports agreement both before (480 of 500) and after (481 of 500). The plan reclassifies as correct the single participant assessed "ask a doctor" by the clinician who self-selected "not right for you." All accuracy measures derive from the pre-mitigation cross-tabulation; the one measure the reclassification touches is negative predictive value, which is reported before (457 of 472) and after (458 of 472) mitigation and labeled accordingly [7].

Two-sided 95% confidence intervals for proportions are Clopper-Pearson exact intervals [29]. The confidence interval for the difference between positive predictive value and sensitivity was obtained by a multinomial bootstrap of the nine cells of the cross-tabulation: 200,000 resamples of 500 participants drawn from the observed cell proportions, with the difference recomputed in each resample and the 2.5th and 97.5th percentiles taken as the interval limits [30]. The random seed, the resample count, and a five-million-resample reference run confirming the interval at one decimal place are recorded in the analysis code.

Supplementary Methods S2. Interval computation and validation for the TACTiC co-primary endpoints

Confidence intervals for the TACTiC endpoints are two-sided 95% Clopper-Pearson exact intervals, the method prespecified in the statistical analysis plan for the co-primary endpoints [24]. As a validation of method, we recomputed the intervals for the endpoints as reported: 1,085 of 1,196 yields 88.9% to 92.3% and 1,132 of 1,154 yields 97.1% to 98.8%, reproducing the published intervals exactly [22]. The unmitigated endpoints were derived by excluding the mitigated participants from each numerator: 1,085 − 78 = 1,007 of 1,196 and 1,132 − 198 = 934 of 1,154. The minimum number of mitigations required for each endpoint to meet its success criterion is the smallest number that, added back to the unmitigated numerator, raises the lower confidence bound above 85%; it is 34 for the first endpoint and 71 for the second. Every quantity in this section is asserted by the analysis script accompanying the manuscript.

Supplementary Methods S3. Decision curve analysis: specification and threshold range

Net benefit was evaluated for three strategies: screening by the CREST application, granting every consumer access without screening, and granting no consumer access. Net benefit was calculated as

where TP is the number of true positives (clinician-eligible consumers approved by the strategy), FP the number of false positives (clinician-ineligible consumers approved), N the total sample size, and pt the threshold probability. For the application, TP = 23 and FP = 3; for unscreened access, TP = 39 and FP = 461; granting no access has zero net benefit at every threshold [7].

In this application, the threshold probability is the probability of guideline eligibility at which taking the product first becomes worthwhile to the consumer; any less confident, and not taking it is the better choice. It is not the ten-year atherosclerotic cardiovascular disease risk threshold used in the treatment guidelines, and the two should not be conflated. The odds of the threshold probability, pt/(1 − pt), function as an exchange rate between the two errors: a threshold of 5% weighs the approval of one eligible consumer as equivalent to 19 inappropriate approvals, and a threshold of 20% as equivalent to 4 [31, 32].

Net benefit was computed across the full range of threshold probabilities, 0% to 100%, and decision curves are displayed across that range (main text, Figure 2). Primary evaluation and interpretation are restricted to a reasonable range of 5% to 20%, specified on clinical grounds, following published recommendations to interpret decision curves only within the range of thresholds a decision maker could plausibly hold [31, 32]. A threshold below 5% would accept more than 19 inappropriate exposures as the price of one eligible consumer treated, a valuation under which screening of any kind serves little purpose. A threshold above 20% would demand better than one-in-five odds of eligibility before a consumer starts a well-tolerated primary prevention therapy, a reluctance out of keeping with the drug’s known safety profile. The bounds thus exclude only valuations under which no screening question arises; within them, the comparison of access strategies is decision-relevant.

The threshold at which the application ceases to outperform granting no access is the threshold at which its net benefit reaches zero, which equals its positive predictive value, 23 of 26 (88.5%). The threshold at which it ceases to outperform unscreened access solves the equality of the two net benefits, 16/474 = 3.4%. Across the reasonable range the net benefit of the application falls from 0.0457 at 5% to 0.0445 at 20%, exceeding both comparators throughout.

Supplementary Methods S4. Prespecification of unmitigated analyses in the TACTiC statistical analysis plan

The statistical analysis plan prespecifies the unmitigated analysis of the first co-primary endpoint twice. In the primary analysis of that endpoint it states that the analysis "will be repeated for the Overall Correct Initial TASS Outcome without any mitigations overall and by subgroups and with a priori mitigations only for the SS population overall and by subgroups and the AUS ITT population overall and by subgroups," and it specifies the same repetition again under Sensitivity Analysis #1 (both at Section 4.2.2, p. 52) [24]. It further specifies a separate table displaying counts and percentages for each mitigation descriptor, distinguishing a priori from post-study mitigations, across the self-selection, actual use intent-to-treat, and per-protocol populations (Section 4.2.2, pp. 52 and 55) [24]. The plan states the intent directly: "The impact of the a priori and post-study mitigations on the overall co-primary endpoints will be assessed by evaluating the endpoints with and without these mitigations" (Section 3.2.3.1, p. 35) [24]. The published supplemental material reports mitigation descriptors for a single population and does not distinguish the two mitigation types [22]. We did not identify the unmitigated analyses in the publication, its supplemental material, the protocol, the statistical analysis plan, or the ClinicalTrials.gov results record [8, 22, 24, 25].

Supplementary Methods S5. Rationale for the two a priori mitigations in the TACTiC statistical analysis plan

Appendix D of the statistical analysis plan (Use and Re-Selection Mitigations, pp. 92–93; clinical study protocol, Table 6, pp. 74–75) states the rationale for the two a priori mitigations in parallel terms: an incorrect affirmative response is mitigated as "a conservative response, which results in them not qualifying for Crestor OTC," a decision to stop treatment as "a conservative action, which results in them not continuing to take Crestor OTC," and in each case the plan concludes that "no medical harm can come to the participant." [8, 24]

Supplementary Methods S6. Endpoint architecture of the TACTiC statistical analysis plan

The endpoint architecture reflects this directly (Figure 2). The statistical analysis plan illustrates the initial self-selection outcomes as a two-by-four table of four clinician outcomes against two participant outcomes, "OK to Use" and "Ask a Doctor," with no participant "Do Not Use" row, and the first co-primary numerator has no term for correct rejection (Section 4.2.2, Figure 1, p. 51) [24]. The final use illustration is a three-by-three table that does include a participant "Do Not Use" row, and the second co-primary numerator accordingly includes a correct rejector term

(Section 4.2.2, Figure 2, p. 53) [24]. Correct rejection is measured where the analysis population permits it and absent where it does not.
